# Off-body Sleep Analysis for Predicting Adverse Behavior in Individuals with Autism Spectrum Disorder

**DOI:** 10.1101/2024.01.23.24301681

**Authors:** Yashar Kiarashi, Pradyumna B Suresha, Ali Bahrami Rad, Matthew A Reyna, Conor Anderson, Jenny Foster, Johanna Lantz, Tania Villavicencio, Theresa Hamlin, Gari D Clifford

## Abstract

Poor sleep quality in Autism Spectrum Disorder (ASD) individuals is linked to severe daytime behaviors. This study explores the relationship between a prior night’s sleep structure and its predictive power for next-day behavior in ASD individuals. The motion was extracted using a low-cost near-infrared camera in a privacy-preserving way. Over two years, we recorded overnight data from 14 individuals, spanning over 2,000 nights, and tracked challenging daytime behaviors, including aggression, self-injury, and disruption. We developed an ensemble machine learning algorithm to predict next-day behavior in the morning and the afternoon. Our findings indicate that sleep quality is a more reliable predictor of morning behavior than afternoon behavior the next day. The proposed model attained an accuracy of 74% and a F1 score of 0.74 in target-sensitive tasks and 67% accuracy and 0.69 F1 score in target-insensitive tasks. For 7 of the 14, better-than-chance balanced accuracy was obtained (p-value<0.05), with 3 showing significant trends (p-value<0.1). These results suggest off-body, privacy-preserving sleep monitoring as a viable method for predicting next-day adverse behavior in ASD individuals, with the potential for behavioral intervention and enhanced care in social and learning settings.

## 1 Introduction

Autism Spectrum Disorder (ASD) is a diverse group of conditions affecting at least 1% of the world’s population [12]. Struggles with social interaction, recurring behaviors, and issues with speech and nonverbal communication characterize it. These individuals’ cognitive and intellectual abilities can vary drastically, thus leading to differing degrees of support needed in their daily routines [10]. Furthermore, ASD often co-occurs with other disorders, with sleep disruption being a significant concern in both children [36, 41, 4] and youth [18, 38, 40]. This issue affects about 40-80% of people with ASD, compared to 10-40% of people without ASD [6, 9, 45].

Previous studies suggested that poor sleep quality can have detrimental effects on brain maturation [17], biological energy transfer [32], memory consolidation [25] and neurobehavioral functioning [46]. This issue is further complicated in individuals with ASD [42, 3, 1, 31]. These studies also revealed that individuals with severe forms of autism often experience more frequent and persistent disruptions in their sleep-wake cycles compared to their less impacted counterparts [48, 50]. Another study, showed that children who slept fewer hours per night had lower verbal skills, adaptive functioning, socialization, and communication skills [53].

Research into the relationship between sleep and challenging behaviors in ASD can be categorized into subjective and objective methodologies. Subjective methodologies employ tools such as sleep logs, diaries [30], and questionnaires [39, 49]. In contrast, objective methodologies utilize technological instruments like polysomnography (PSG) [33], actigraphy [47, 35], wearable devices, and mobile applications [55, 29, 34].

While subjective methods provide first-hand accounts of sleep patterns and behaviors, thereby offering valuable insights into personal experiences, they are subject to personal biases and recall limitations, potentially compromising their reliability. On the other hand, objective methods furnish a more concrete, quantifiable measure of sleep quality, yielding data-driven insights less susceptible to individual interpretation. However, these objective techniques often require the use of on-body sensors or wearable devices, which may result in discomfort that could adversely affect sleep quality. Additionally, on-body sensors may not be well-tolerated in those with ASD who present with sensory sensitivities. As the result, typically, these two approaches were employed in conjunction for more comprehensive results [43, 22, 37].

Findings across these studies consistently suggested a correlation between sleep quality and behavioral issues. They revealed that poor sleep is associated with more restricted and repetitive behaviors [22], showed improvements in daytime behavior following sleep interventions [37], indicated that sleep variables account for variances in daytime functioning [43], highlighted that individuals with severe sleep issues display higher levels of challenging behaviors [2], and even proposed that the severity of ASD can be predicted based on the hours of sleep per night [49].

Although research established that children with ASD often face severe sleep issues that exacerbate their daytime behavioral challenges, such as aggression, disruptive behavior, and self-injurious behaviors [28, 23, 13], the prospect of predicting next-day behavior using sleep studies remained largely unexplored in both academic literature and clinical guidelines. Existing treatment protocols for managing ASD-related behaviors frequently either neglected sleep issues or provided only a cursory overview. One factor contributing to this research and guideline gap could be the limitations inherent in current sleep assessment tools, which may struggle to accurately capture the nuanced sleep disturbances commonly observed in both children and adults with ASD. Recent research has demonstrated the possibility of predicting next-day adverse behaviors using night-to-night variation in sleep timing and duration in individuals with ASD [14]. The study used support vector machine (SVM) classifiers to analyze an extensive dataset of over 20,000 nightly sleep observations matched with subsequent daytime behaviors. Sleep metrics such as total sleep time and sleep efficiency were utilized to gauge sleep quality. The study established 58%-60% balanced accuracy for different target behaviors across individuals and achieved a statistically significant predictive relationship in 81% of the subjects. A key limitation is the method’s heavy reliance on caretaker observations, which require monitoring the individuals at 15-minute intervals throughout the night, thereby introducing potential observer bias and raising concerns about scalability.

More recently, studies have shown the potential of bed sensors as a more feasible solution for predicting daytime behaviors in children with ASD [5]. The study achieved accuracies of 78% and 79% for predicting daytime adverse behavior using SVM and artificial neural network (ANN) classifiers, respectively. However, the study was based on a sample size of just two individuals with ASD. Given the heterogeneous nature of ASD, where symptoms and responses can vary significantly among individuals, such a limited sample size precludes drawing any generalizable conclusions. Moreover, bed sensors come with inherent limitations. They are typically placed under the mattress, which can result in high false positive rates[44], as they may be easily affected by other movements or environmental factors.

The disproportionate focus on individuals with mild forms of autism in sleep research overlooks a critical demographic, those with more severe and complex presentations [13], who often experience the most severe sleep disruption and behavioral problems that remain under-explored. Conventional sleep assessment tools like actigraphy and polysomnography often hinge on the individual’s ability to communicate and cooperate, limiting their applicability in cases of severe autism. This research gap signifies an urgent need for developing alternative sleep monitoring approaches that are tailored to individuals with severe autism. Such approaches should minimize the reliance on the individual’s communication skills or the constant presence of a care partner, thereby providing a more inclusive and accurate understanding of sleep disturbances across the autism spectrum.

In our study, utilizing a low-cost near-infrared camera, we extracted motion data in a privacy-preserving manner, computing statistics at the edge to assess sleep quality in individuals with severe autism. Over more than a year, we gathered data that offers a broader context for understanding sleep patterns and related behaviors in ASD. This longitudinal dataset, including overnight movement signals and behavioral labels, aids in analyzing the relationship between sleep quality and daytime behaviors such as self-injurious behavior, aggression, and disruptive behavior. Our proposed model was formed by an ensemble of seven machine learning models to create a robust model to predict the next day’s behavior in the morning and the afternoon using 1-7 previous sleep data.

## 2 Data Collection

### 2.1 Participants

The study took place at The Center for Discovery (TCFD) in New York State, which provides educational, medical, clinical, and residential care to individuals with severe and complex disabilities including ASD. The participants in this study required a residential level of care having been unsuccessful in a less restrictive environment due to the severity and complexity of their conditions. The center has an internal Institutional Review Board that reviewed and approved of this study protocol.

Fourteen individuals previously diagnosed with ASD ages 15 to 22 (Mean ± Standard Deviation (SD): 18.23 ± 1.84) participated in this study. The sample consisted of 11 white (accounting for 78.6%), two Asian (14.2%) and one African American (7.1%). In terms of gender, 12 were males (85.7%) and 2 were females (14.2%).

The Autism Spectrum Rating Scales (ASRS) [24] is a standardized, norm-referenced instrument that measures behaviors associated with ASD and aligns with criteria from the *Diagnostic and Statistical Manual of Mental Disorders*, Fifth Edition (DSM-5) [51]. Because the ASRS is not normed for participants over the age of 18, the last administered ASRS was included for those over 18 during the data collection period. T-scores from 60-64 are in the Slightly Elevated classification; 65-69 Elevated classification; 70-85 Very Elevated classification. As referenced in Table 1, the number and pattern of symptoms associated with ASD as measured by the ASRS DSM-5 Scale was in the Elevated range for three participants and Very Elevated for 11. All participants met DSM-5 criteria for a Level 1 or Level 2 severity for ASD, requiring “substantial” to “very substantial” support based on clinical judgment by a licensed psychologist.

**Table 1:**
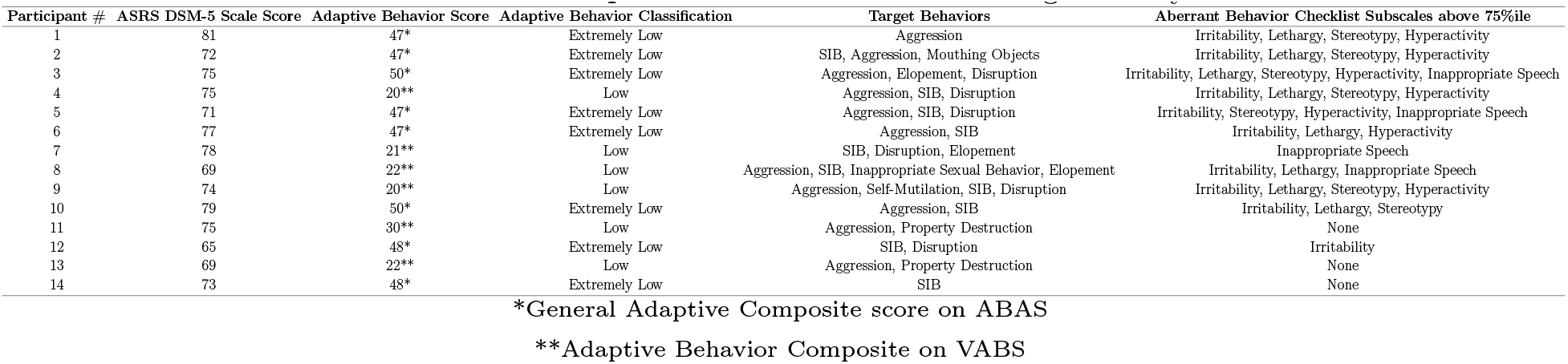
Participant Characteristics Indicating Severity.

**Table 2:** Performance Metrics for Target-sensitive and Target-insensitive Models During Morning and Afternoon.

| Prediction Task | Time | Precision | Recall | F1 Score | Accuracy |
| --- | --- | --- | --- | --- | --- |
| Target-Sensitive | Morning | $0.73 \pm 0.34$ | $0.75 \pm 0.39$ | $0.74 \pm 0.35$ | $0.74 \pm 0.15$ |
| Target-Insensitive | Morning | $0.61 \pm 0.17$ | $0.80 \pm 0.34$ | $0.69 \pm 0.21$ | $0.67 \pm 0.15$ |
| Target-Sensitive | Afternoon | $0.39 \pm 0.26$ | $0.11 \pm 0.09$ | $0.17 \pm 0.13$ | $0.74 \pm 0.18$ |
| Target-Insensitive | Afternoon | $0.44 \pm 0.3$ | $0.16 \pm 0.27$ | $0.23 \pm 0.19$ | $0.75 \pm 0.21$ |

In most cases, cognitive testing was not able to be conducted due to language limitations and inability to respond to test items. As such, level of intellectual disability was determined based on scores on adaptive behavior measures. As part of standard practice, TCFD school psychologists administer either the Adaptive Behavior Assessment System (ABAS) [27] or Vineland Adaptive Behavior Scale-3rd Edition (VABS-3) [26]. The ABAS yields a “General Adaptive Composite” (GAC) score that represents a norm-referenced score for the individual. The VABS-3 provides an “Adaptive Behavior Composite” norm-referenced score based on five domains measured. As can be seen in Table 1, all participants functioned in the moderate or severe range of intellectual disability with adaptive scores in the lowest classification range designated for the particular measure (Low for the VABS-3 and Extremely Low for the ABAS).

Participants had behavior intervention plans to address target behaviors that interfered with functioning. For the purposes of this study, only aggression (e.g., hitting, biting, kicking, scratching), self-injurious behaviors (SIB) (e.g., hitting body parts on objects or with hands, biting self) and disruption (e.g., screaming, throwing objects, dropping) were included in the analysis given their frequency in the sample. The Aberrant Behavior Checklist [8] is a measurement tool that assesses maladaptive behaviors in children and adults with intellectual disabilities. It yields scores in five areas: Irritability/Agitation, Lethargy/Social Withdrawal, Stereotypic Behavior, Hyperactivity/Noncompliance, and Inappropriate Speech. As shown in Table 1, 11 of the participants had at least one subscale on the ABC that was above the 75th percentile, indicating more extreme behavioral presentations compared to a normative sample of peers with disabilities in community-based special education programs [7].

### 2.2 Data sources

Board Certified Behavior Analysts (BCBAs) at TCFD prescribe and implement BIPs, and behavior data for the challenging behaviors is collected continuously by trained direct care staff across three shifts in a 24-hour period: morning (7:00-3:00) afternoon (3:00-23:00), and overnight (23:00-7:00). Additionally, Raspberry Pi based sensor units with infrared cameras were installed in each participant’s bedroom to record movement across the evening period (19:00-8:00). To preserve privacy, no images were recorded; movement was captured by comparing and recording changes in light intensity, pixel by pixel, from one frame to the next across the recording period. Overnight sensor data was paired with the corresponding participant’s behavior data for this analysis.

The study was approved by the Emory Institutional Review Board and is registered under the project title “Utilizing Technology to Accurately Track Sleep and Predict Adverse Behaviors in Autism” (IRB00003823).

### 2.3 Sensing pipeline

We employed a Raspberry Pi 4 IR camera (RPi-IR) for data acquisition. The detailed process of movement data acquisition and analysis is illustrated in Figure 1. Our system also incorporates a 5MP OV5647 infrared camera, which has a native resolution of 2592 × 1944 pixels and a 60 Hz frame rate. For computational efficiency and privacy preservation, we downsampled the captured frames to 5 × 4 pixels and down sampled the frame rate to 1.5Hz.

**Figure 1:**
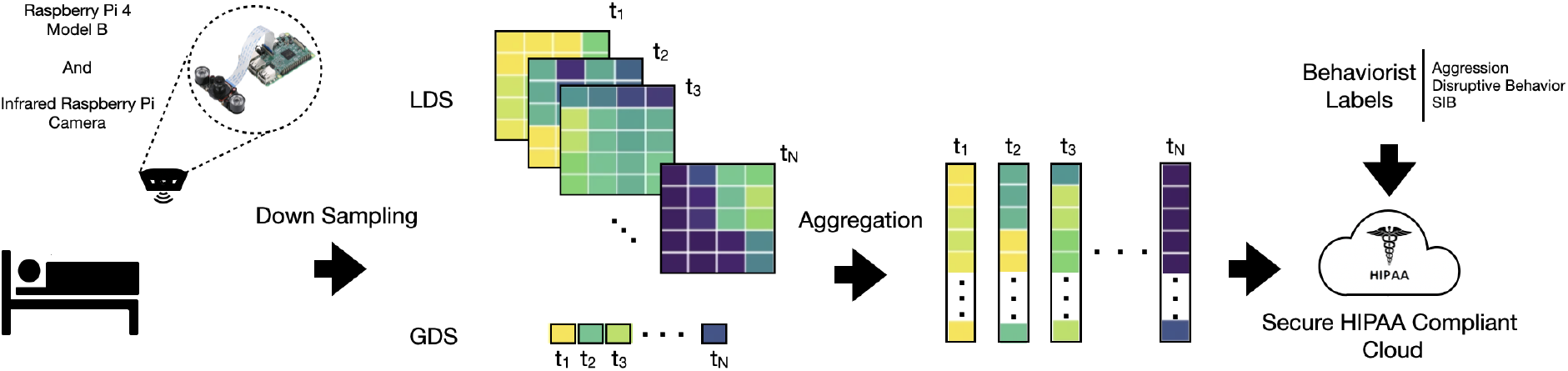
Diagram illustrating the workflow for collecting movement data using a Raspberry Pi 4 Model B equipped with a 5MP OV5647 infrared camera. The camera has a resolution of 2592 × 1944 and a frame rate of 60fps. Frames are downsampled to 5×4 superpixels (with 4 superpixels wide and 5 superpixels high), and the frame rate is resampled to 1.5Hz for analysis. Both Global Difference Sum(GDS) and Local Difference Sum(LDS) are calculated. These features are aggregated for each time frame to generate a multidimensional time series. The resulting data is then securely uploaded to a HIPAA-compliant cloud. In the days following data collection, the behaviorist logs resident labels during two sessions (morning and afternoon) for three distinct target adverse behaviors: aggression, disruptive behavior, and self-injurious behavior (SIB).

## 3 Methods

The objective of this study was to assess the predictive association between movements during prior night of sleep, serving as an indicator of sleep quality, and challenging behavior exhibited during daytime.

### 3.1 Preprocessing

Under certain assumptions, our algorithms calculate both the Global Difference Sum (GDS) and Local Difference Sum (LDS) [52] for each frame at a distinct time *t* with pixel resolution *M* × *N*. The assumptions were as follows:

1. We have only one person in the entire video.
2. The background is static; the only moving object is either the person or an object attached to them.
3. The video is free from corruption and noise.

Given a frame *F*_*t*_ and its preceding frame *F*_*t*−1_, the difference frame *D*_*t*_ is *D*_*t*_ = *F*_*t*_ − *F*_*t*−1_. The GDS is mathematically expressed for each difference frame *D*_*t*_ as:

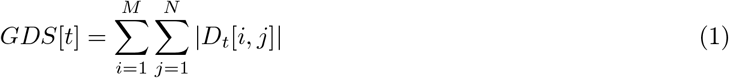

Similarly, for LDS calculations, the difference frame *D*_*t*_ is partitioned into *K* × *L* equally sized local blocks *DL*_*t,s*_, where we set *K* = 5 and *L* = 4. The LDS is then given by:

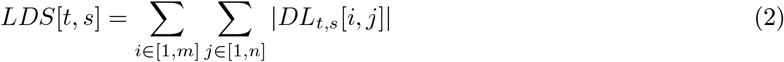

These GDS and LDS values are aggregated over each time frame to produce a multidimensional time series. This resulting data was securely uploaded to a HIPAA-compliant cloud for further analysis. In a parallel process, trained staff record labels for resident behaviors during two separate sessions (i.e., morning and afternoon), focusing on three adverse behaviors: aggression, disruptive behavior, and SIB.

### 3.2 Feature extraction

The methodology employed in this study is schematically depicted in Figure 2, feature extraction is the initial phase of our methodological approach. We extract five features for each dimension-modality (i.e., 20 for GDS and one for LDS). This process generates a 5-dimensional feature vector for GDS signals and a 100-dimensional feature vector for LDS signals, culminating in a combined 105-dimensional feature vector. The five crafted features are as follows:

**Figure 2:**
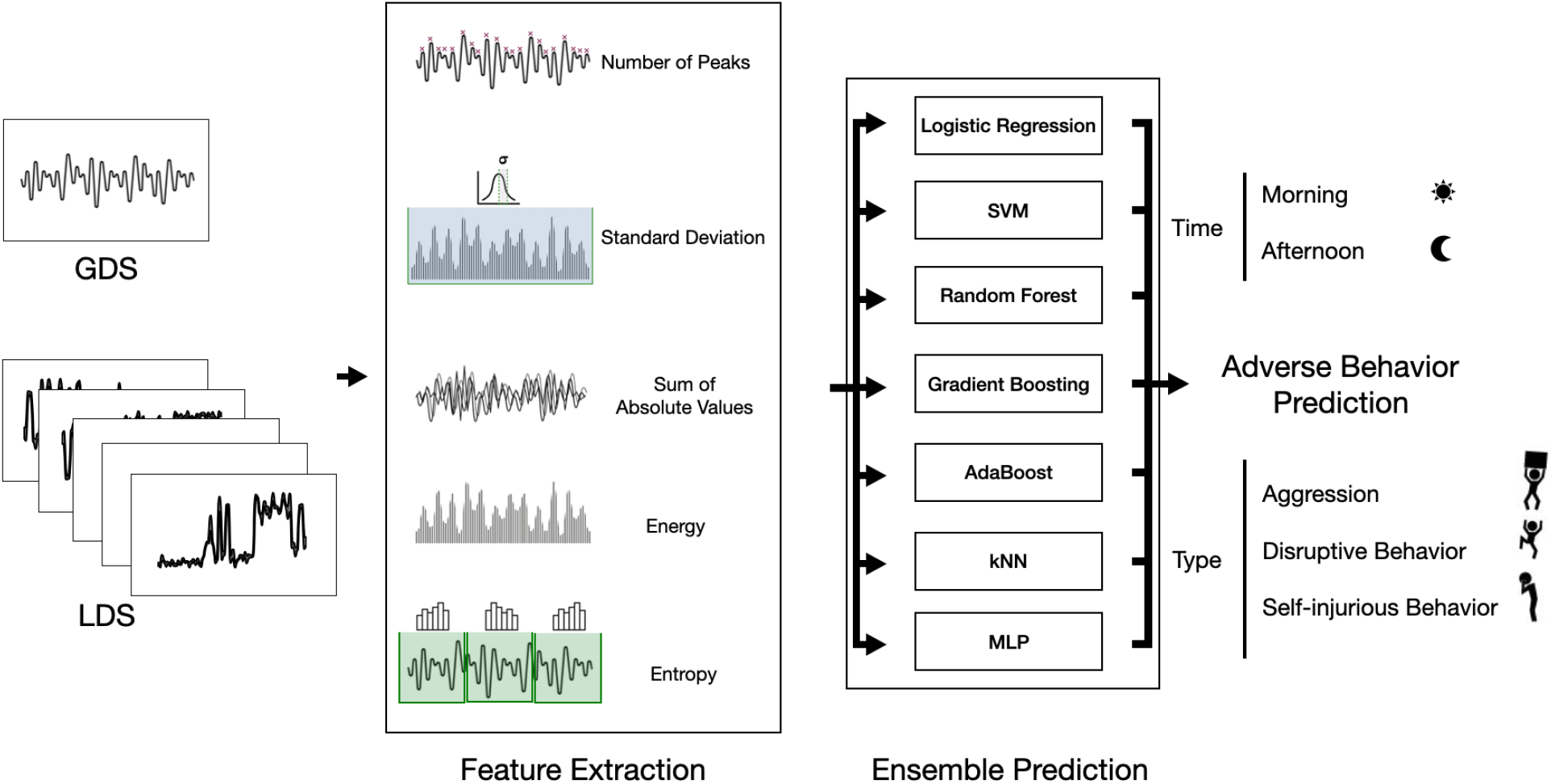
Flowchart illustrating the two-stage methodological approach for sleep analysis. The first stage involves feature extraction from Global Difference Sum (GDS) and Local Difference Sum (LDS), resulting in a 105-dimensional feature vector for each observation. Five specific features are extracted, each chosen for its relevance in sleep analysis. The second stage employs an ensemble of seven machine learning algorithms to make predictive assessments based on the extracted features.

1. **1-norm (Sum of Absolute Values)**: This measure captures the overall fluctuations within the data series and can be indicative of restless sleep or sleep quality. Larger values may signify frequent movements or disruptions.

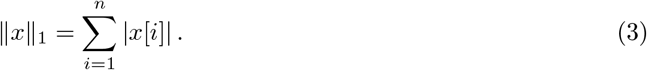 Here, *x*[*i*] represents a single dimension of either GDS or LDS in the *i*th time stamp. Let *n* denote the number of timestamps recorded in a single night for a resident.
2. **Standard Deviation (***s*): Standard deviation gauges the variability in the data, which can be useful for detecting anomalies in sleep patterns.

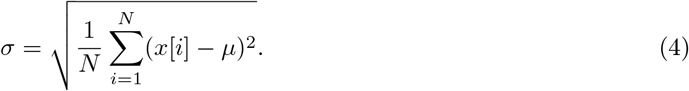 Here, *μ* is the mean of *x* and *N* (see Fig 1) is the total length of the signal over a night.
3. **Number of Peaks (NumPeaks)**: Peaks in the data can represent moments of heightened activity or agitation, which could be of relevance in diagnosing sleep disorders like insomnia or sleep apnea.

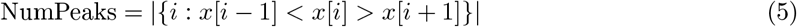
4. **2-norm (Energy)**: 2-norm provides an overall sense of changes in consecutive frames. In this context, a high-energy signal can indicate restless or disrupted sleep.

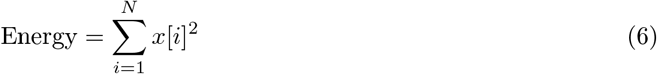
5. **Entropy (***H*_**global**_): To quantify the complexity or unpredictability of sleep, we calculate the global entropy (*H*_global_). Higher entropy values might suggest irregular sleep patterns. The global entropy is computed as an average of local entropies across the signal.

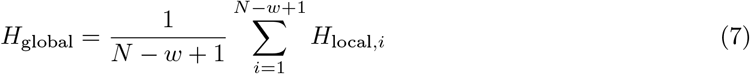 In this equation, *w* is the window size used to calculate each local entropy (*H*_local,*i*_). Specifically, for each window of size *w* starting at index *i* in the time series *x*[*i*], we calculate a local entropy using the following formula:

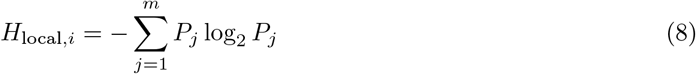

where *P*_*j*_ represents the empirical probability of the *j*-th bin in a histogram constructed from the windowed segment of the signal, spanning from *x*[*i*] to *x*[*i* + *w* − 1]. This histogram is divided into *m* bins. The empirical probability *P*_*j*_ is calculated as the proportion of samples falling into the *j*-th bin within the window, thus defining an empirical probability distribution based on the observed values of *x*[*i*] over each window. In this study, we set *w*=100.

### 3.3 Prediction Models

Upon extracting features mentioned in Section 3.2, seven predictive models were applied as depicted in Figure 2. These algorithms include the support vector machine (SVM) [15], logistic regression [16], random forests [11], gradient boosting [20], AdaBoost [19], *k*-nearest neighbors (kNN), and multilayer perceptrons (MLP) [21]. Finally, a voting algorithm consolidates the predictive outcomes from all seven models, enhancing the system’s robustness and reliability by opting for the majority vote as the final prediction label.

Each model is trained using the 105-dimensional aggregated feature vector for prediction tasks, such as forecasting aggression. We repeated this analysis for morning and afternoon for:

1. **Target-sensitive Prediction:** In this study, the predictive models were designed to forecast both the occurrence and timing of specific forms of adverse behavior at designated times of day. For instance, a single data instance might be labeled with multiple behaviors such as aggression and self-injurious behaviors. The model aims to accurately predict the absence of a particular adverse behavior even when other adverse behaviors are concurrently present.
2. **Target-insensitive Prediction:** Here, the focus was on predicting the time and presence of any adverse behavior, regardless of its specific type, at a particular time of day.

To develop these models, we allocated 80% of the data for training purposes and reserved the most recent 20% to preserve temporal causality in each subject-specific scenario, such as forecasting aggressive behavior by a specific resident the following day.

### 3.4 Evaluation metrics

To assess the efficacy of our model, we utilized both conventional evaluation metrics and significance testing against a prevalence-aware baseline.

#### 3.4.1 Conventional evaluation metrics

We evaluated the proposed model using conventional metrics commonly employed for predictive models. These include accuracy, which provides a general measure of model performance; precision, which gauges the model’s ability to identify positive cases correctly; recall, which measures the model’s ability to capture all potential positive cases; and F1 score, which provides a balanced measure that takes into account both precision and recall. All metrics were derived through micro-averaging, which combine the individual counts of True Positives, False Positives, and False Negatives across all classification categories to formulate a consolidated measure. The benefit of utilizing micro-averaging lies in its sensitivity to the performance of each instance, thereby giving more weight to the classes that are more populous in the dataset. For robust statistical representation, all evaluation outcomes are conveyed as Mean±Standard Deviation over individuals. These metrics serve as an initial yardstick for evaluating the effectiveness of our predictive models.

#### 3.4.2 Validation Through Permutation Testing

To assess the statistical significance of our model’s predictive accuracy beyond mere random chance, we employed a permutation-based significance testing approach. This method involved reshuffling the test set labels to create a distribution of accuracy scores under the null hypothesis, where the model’s performance is assumed to be no better than random guessing. Through this process, we were able to compare the actual model accuracy against this distribution to ascertain whether the observed performance was statistically significant. The approach is detailed in Algorithm 1.

The null hypothesis, *H*_01_, in this test, suggests that the observed accuracy is not superior to that achievable by random chance, given the label distribution.

##### Algorithm 1 Permutation Test

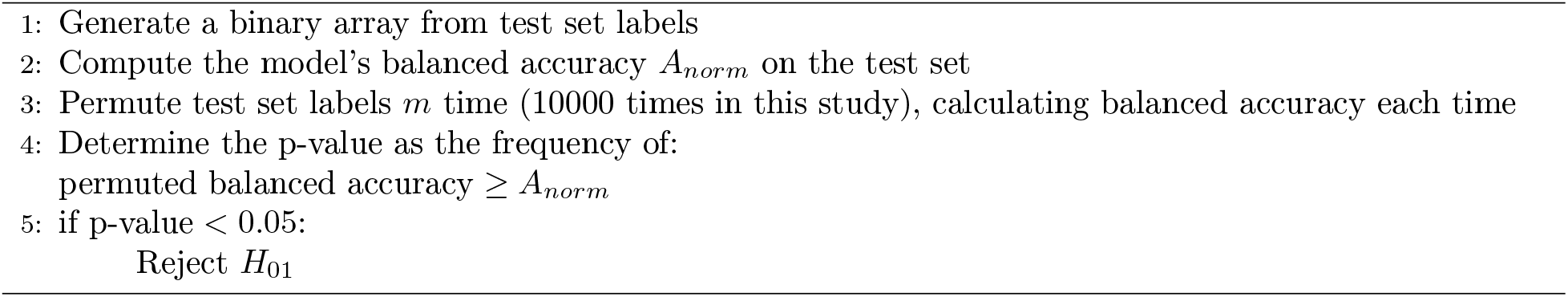

### 3.5 Assessment of Temporal Sleep-Behavior Patterns

We explored the impact of incorporating data from previous nights to improve the predictive accuracy of our models. This enhancement was accomplished by progressively expanding the input feature set, incorporating data from 1 to 7 preceding nights, in addition to the original features obtained from a single night’s data. To achieve this, we concatenated the feature vectors from consecutive nights, thereby forming an extended feature set for each subsequent night. Subsequently, we applied the model introduced in Section 3.3 to these enhanced feature sets to predict the day time behavior.

### 3.6 Evaluation of Predictive Models

To evaluate the role of each AI-based model presented in Section 3.3. We trained and evaluated each model with the composite 105-dimensional feature vector (single night), derived from sleep data. The training procedure for these models involved allocating 80% of the data for training, while the remaining 20% was set aside for testing purposes. This approach was adopted to maintain the temporal causality essential for predicting subject-specific behaviors, as detailed in Section 3.3.

## 4 Results

### 4.1 Performance Analysis

Table 5 and Fig 3 show the morning and afternoon session performance metrics for each prediction task. We computed the standard deviation and the mean balanced accuracy, presenting the results as mean accuracy (MA)± standard deviation (SD).

**Figure 3:**
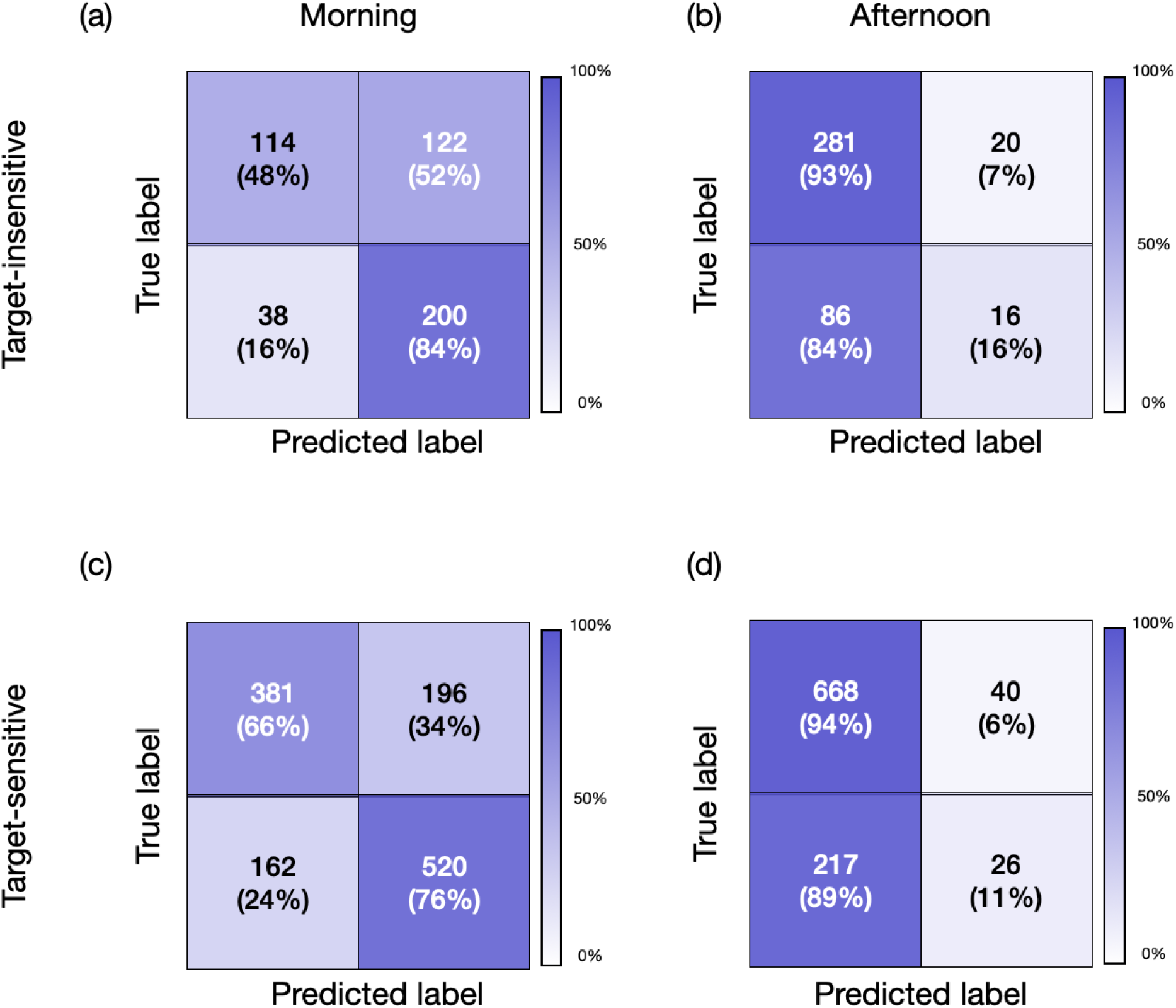
Confusion matrices for target-sensitive and target-insensitive models predicting the presence of adverse behavior at different times of the day: (a) Morning using the target-sensitive model, (b) Afternoon using the target-sensitive model, (c) Morning using the target-insensitive model, and (d) Afternoon using the target-insensitive model.

Table 5 presents model performance differences. In the morning, the target-sensitive model had a macro average accuracy of 0.74 ± 0.15, compared to 0.67 ± 0.15 for the target-insensitive model. In the afternoon, these accuracies changed to 0.74 ± 0.18 and 0.75 ± 0.21 for the target-sensitive and insensitive models. However, the afternoon F1 scores were 0.17 ± 0.13 and 0.23 ± 0.19, indicating the inflated accuracy for predicting the presence of adverse behavior in the afternoon.

Fig. 3 further illustrates the challenge in accurately predicting the presence of adverse behavior in the afternoon sessions. The confusion matrices for the morning, shown in Fig. 3 panels (a) and (c), reveal that the target-sensitive model achieved a micro balanced accuracy of 71%, compared to 66% for the target-insensitive model. Conversely, the afternoon session, depicted in panels (b) and (d) of Fig. 3, demonstrates a decrease in balanced accuracy to 54.5% for the target-sensitive model and 52.5% for the target-insensitive model.

Building on the earlier results of model performance in predicting adverse behavior, Table 3 and Figure 4 offer an in-depth look at how these models perform in identifying specific types of adverse behaviors during morning sessions. For aggression, self-injurious behavior, and disruptive behavior, the models achieved macro average balanced accuracies of 0.75 ± 0.19, 0.75 ± 0.33, and 0.76 ± 0.17 respectively. The corresponding macro average F1 scores were 0.74 ± 0.38, 0.74 ± 0.44, and 0.76 ± 0.4. In terms of micro average balanced accuracies, the values were 0.70 for aggression, 0.73 for self-injurious behavior, and 0.72 for disruptive behavior, accompanied by micro average F1 scores of 0.68, 0.8, and 0.75, respectively. These findings highlight the models’ overall effectiveness in predicting different adverse behaviors.

**Table 3:** Performance Metrics for Different Adverse Behaviors in Morning.

| Adverse Behavior | Precision | Recall | F1 Score | Accuracy |
| --- | --- | --- | --- | --- |
| Aggression | $0.73 \pm 0.38$ | $0.75 \pm 0.44$ | $0.74 \pm 0.38$ | $0.75 \pm 0.19$ |
| SIB | $0.68 \pm 0.41$ | $0.82 \pm 0.5$ | $0.74 \pm 0.44$ | $0.75 \pm 0.33$ |
| Disruptive Behavior | $0.8 \pm 0.37$ | $0.73 \pm 0.43$ | $0.76 \pm 0.4$ | $0.76 \pm 0.17$ |

**Figure 4:**
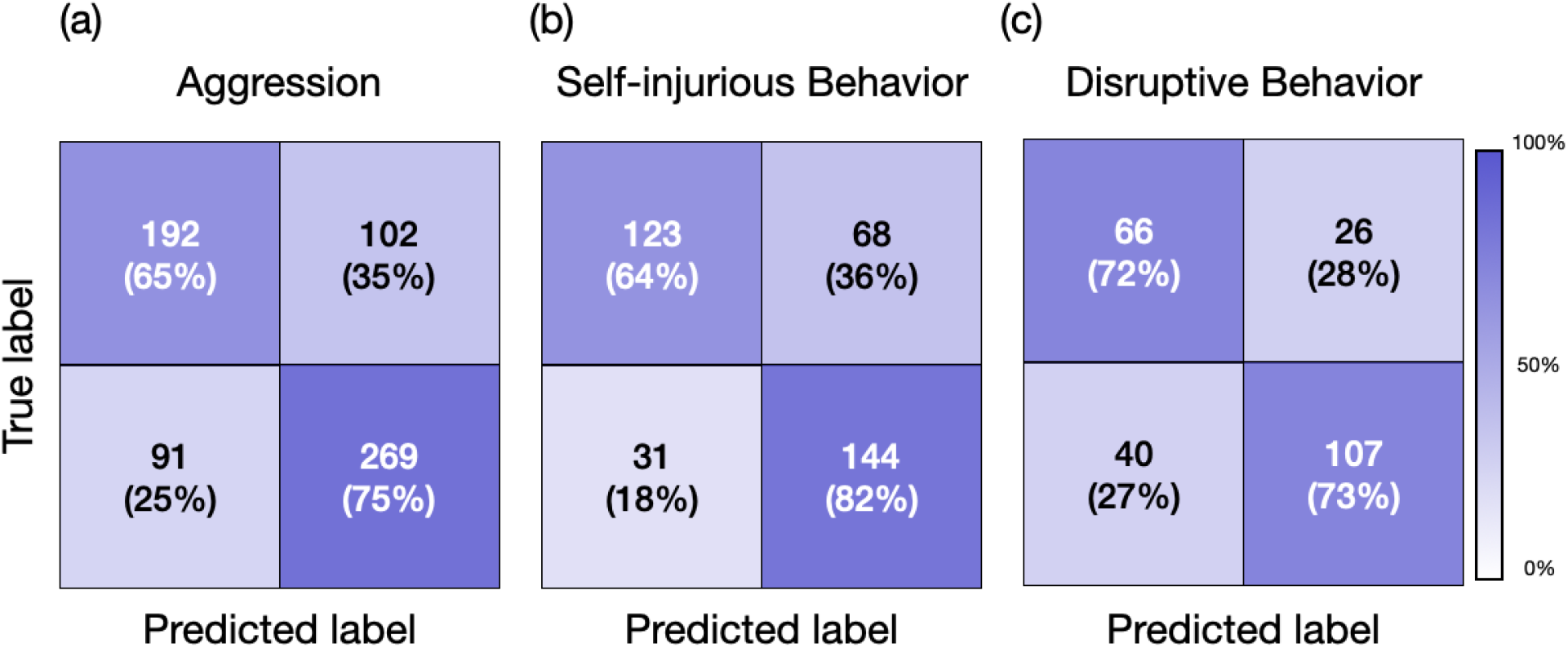
Confusion matrices for predicting different adverse behaviors in the morning time. Confusion matrix for predicting (a) Aggression, (b) Self-injurious behavior, and (c) Disruptive behavior.

### 4.2 Significance Analysis

Following the evaluation of our model, we carried out the significance tests outlined in Section 3.4.2. These tests were applied to each resident for various adverse behaviors. The resulting p-values are shown in Table 4. This table highlights p-values falling below the 0.05 threshold in bold, denoting instances where the null hypothesis can be rejected. This suggests significant findings in contrast to the permutation test detailed in Section 3.4.2). For scenarios where data points are absent for specific resident-task combinations, “N/A” is indicated.

**Table 4:**
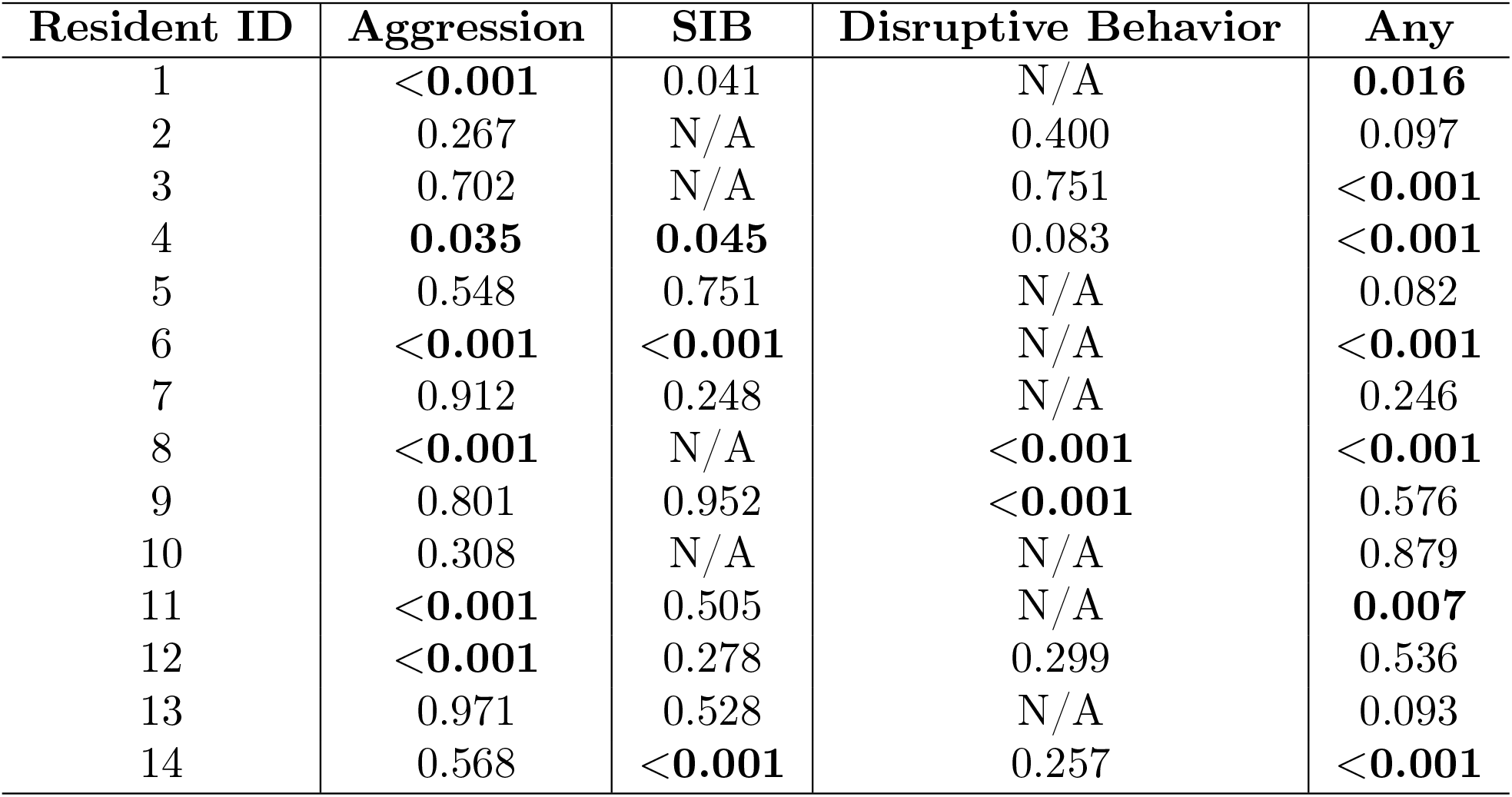
Significance Testing Across Different Residents (p-values)

| Resident ID | Aggression | SIB | Disruptive Behavior | Any |
| --- | --- | --- | --- | --- |
| 1 | <0.001 | 0.041 | N/A | **0.016** |
| 2 | 0.267 | N/A | 0.400 | 0.097 |
| 3 | 0.702 | N/A | 0.751 | <0.001 |
| 4 | **0.035** | **0.045** | 0.083 | <0.001 |
| 5 | 0.548 | 0.751 | N/A | 0.082 |
| 6 | <0.001 | <0.001 | N/A | <0.001 |
| 7 | 0.912 | 0.248 | N/A | 0.246 |
| 8 | <0.001 | N/A | <0.001 | <0.001 |
| 9 | 0.801 | 0.952 | <0.001 | 0.576 |
| 10 | 0.308 | N/A | N/A | 0.879 |
| 11 | <0.001 | 0.505 | N/A | **0.007** |
| 12 | <0.001 | 0.278 | 0.299 | 0.536 |
| 13 | 0.971 | 0.528 | N/A | 0.093 |
| 14 | 0.568 | <0.001 | 0.257 | <0.001 |

**Table 5:** Metrics for Target-Insensitive and Target-Sensitive Models Across Different Models in Morning.

| **Prediction Approach** | **Model** | **Precision** | **Recall** | **F1 Score** | **Accuracy** |
| --- | --- | --- | --- | --- | --- |
| Target-Insensitive | AdaBoost | $0.6 \pm 0.3$ | $0.63 \pm 0.35$ | $0.61 \pm 0.3$ | $0.59 \pm 0.19$ |
| Target-Insensitive | Logistic Regression | $0.58 \pm 0.28$ | $0.72 \pm 0.27$ | $0.65 \pm 0.26$ | $0.59 \pm 0.17$ |
| Target-Insensitive | MLP | $0.57 \pm 0.31$ | $0.62 \pm 0.41$ | $0.6 \pm 0.32$ | $0.57 \pm 0.22$ |
| Target-Insensitive | Random Forest | $0.62 \pm 0.32$ | $0.81 \pm 0.4$ | $0.7 \pm 0.32$ | $0.64 \pm 0.23$ |
| Target-Insensitive | SVM | $0.61 \pm 0.33$ | $0.92 \pm 0.49$ | $0.73 \pm 0.38$ | $0.65 \pm 0.19$ |
| Target-Insensitive | XGBoost | $0.63 \pm 0.27$ | $0.71 \pm 0.36$ | $0.67 \pm 0.29$ | $0.62 \pm 0.15$ |
| Target-Insensitive | kNN | $0.62 \pm 0.33$ | $0.8 \pm 0.42$ | $0.7 \pm 0.36$ | $0.64 \pm 0.2$ |
| Target-Insensitive | Voting | $0.61 \pm 0.17$ | $0.80 \pm 0.34$ | $0.69 \pm 0.21$ | $0.67 \pm 0.15$ |
| Target-Sensitive | AdaBoost | $0.69 \pm 0.32$ | $0.56 \pm 0.27$ | $0.62 \pm 0.27$ | $0.63 \pm 0.14$ |
| Target-Sensitive | Logistic Regression | $0.67 \pm 0.28$ | $0.64 \pm 0.29$ | $0.66 \pm 0.27$ | $0.64 \pm 0.12$ |
| Target-Sensitive | MLP | $0.68 \pm 0.25$ | $0.73 \pm 0.29$ | $0.71 \pm 0.25$ | $0.67 \pm 0.12$ |
| Target-Sensitive | Random Forest | $0.74 \pm 0.34$ | $0.65 \pm 0.33$ | $0.7 \pm 0.32$ | $0.687 \pm 0.13$ |
| Target-Sensitive | SVM | $0.7 \pm 0.29$ | $0.82 \pm 0.36$ | $0.76 \pm 0.29$ | $0.71 \pm 0.15$ |
| Target-Sensitive | XGBoost | $0.77 \pm 0.25$ | $0.58 \pm 0.28$ | $0.66 \pm 0.24$ | $0.69 \pm 0.15$ |
| Target-Sensitive | kNN | $0.79 \pm 0.32$ | $0.77 \pm 0.37$ | $0.74 \pm 0.34$ | $0.7 \pm 0.18$ |
| Target-Sensitive | Voting | $0.73 \pm 0.34$ | $0.76 \pm 0.39$ | $0.75 \pm 0.35$ | $0.74 \pm 0.15$ |

In predicting the type of adverse behaviors in the morning, the null hypothesis was rejected for aggression predictions in 6 out of 14 instances. For SIB, significant outcomes were observed in 3 out of 10 cases. In the case of disruptive behavior, the permutation test indicated significance in 2 out of 7 instances. Additionally, the permutation test contradicted the null hypothesis for predictions involving target-insensitive adverse behaviors in 7 out of 14 scenarios.

### 4.3 Predictive Value of Prior Nights

Figure 5, panels (a) and (b), illustrate the models’ accuracy and F1 score, respectively. The findings reveal a modest enhancement in performance with the incorporation of multiple nights’ data. Specifically, the average accuracy for the residents improved from 0.67 (using data from previous night) to 0.68 when data from 7 prior nights was used. A more pronounced improvement was observed in the mean F1 score, which increased from 0.69 (using data from previous night) to 0.74 with the inclusion of data from 7 preceding nights.

**Figure 5:**
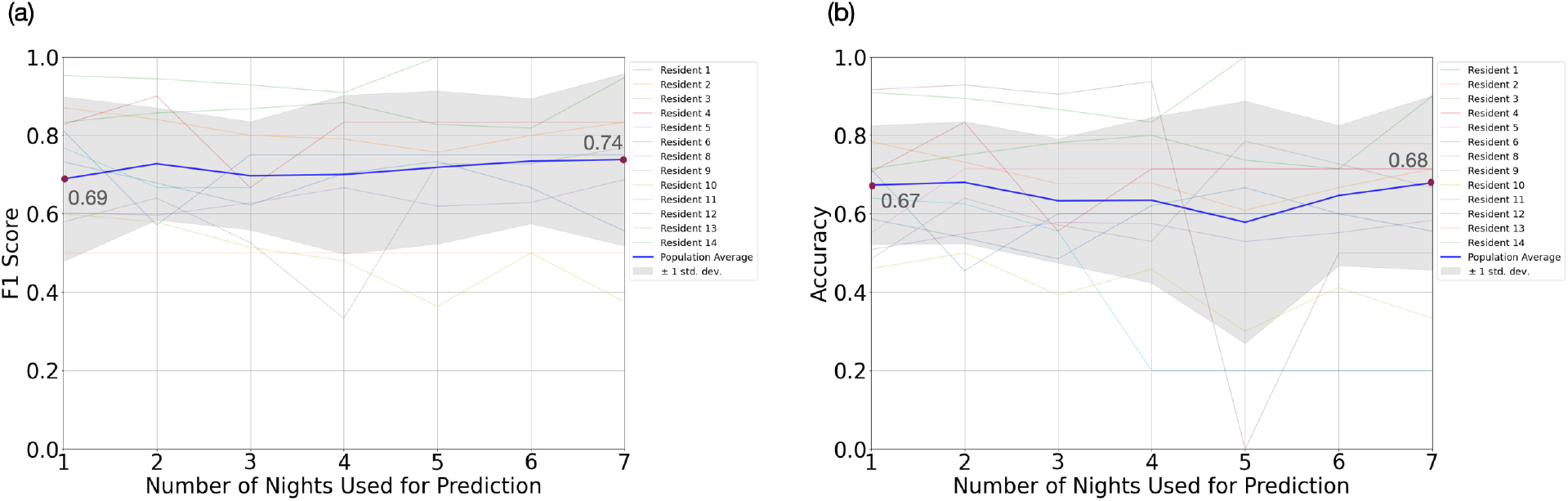
Performance analysis of predictive models using different lengths of historical nights’ data(a) F1 Score and (b) Accuracy across different residents using various lengths of consecutive nights for prediction. The graphs showcase the performance outcomes, represented by F1 score and accuracy, for different residents when using varying lengths of consecutive nights’ data for prediction. It is notable that utilizing data from a 7-day period yields the highest results for both metrics, even slightly surpassing the performance achieved when only data from a single preceding night is used.

### 4.4 Model Assessment

The Ensemble Voting model achieved an F1 score of 0.69 ± 0.21 and an accuracy of 0.67 ± 0.15. This performance notably exceeds AdaBoost, Logistic Regression, and MLP. However, while Random Forest, kNN, and SVM recorded F1 scores of 0.7 ± 0.32, 0.7 ± 0.36, and 0.73 ± 0.38 respectively, the increased standard deviations in their Accuracy and Recall metrics indicate less consistency and reliability. Our Ensemble Voting model offers an advantageous balance, providing robust performance and reliability across various metrics in the target-insensitive model category.

For the target-insensitive task, we observed comparable performance across different metrics for both the SVM and Voting algorithms. These algorithms outperformed the remaining models, achieving an accuracy of 0.71 ± 0.15. Furthermore, the F1 scores registered were 0.76 ± 0.29 for the SVM and 0.75 ± 0.35 for the voting model.

## 5 Discussion

The findings of this study suggest the potential for predicting next-day adverse behaviors in individuals with ASD by monitoring the previous night’s activities using a non-invasive, off-body sensor. This approach could offer a valuable daily feedback mechanism for caregivers and healthcare professionals, enabling them to optimize resource allocation and intervention strategies based on the monitored data. The utilization of such a system promises to enhance the understanding and management of behavior in the ASD population.

The proposed model demonstrated acceptable predictive performance, evidenced by its statistical significance in the permutation test (Section 3.4.2) for 7 out of 14 residents. Furthermore, the model indicated a trend towards significance for 3 out of the 14 residents, with a p-value lower than 0.1, as detailed in Table 4. To quantify the effect size, we calculated the Cohen’s *d* coefficient, obtaining a value of 0.45 ± 0.18. This indicates a medium effect size and suggests that while the prediction model did not uniformly predict adverse behavior across the entire population, it was notably effective in a substantial subset. This partial efficacy might be attributable to various factors such as differences in medication (e.g., melatonin usage), distinct sleep movement patterns in the ASD population (where certain adverse behaviors may not be detectable via movement sensors and might require more invasive methods like Polysomnography for identification), changes in the distribution of adverse behavior during the training and testing phase (20% most recent nights) or limitations in the scalability of the model to the entire population.

As presented in Table, there is a significant difference in model performance between morning and afternoon sessions, which deserves consideration, given that both models utilize data from the previous night’s sleep for their predictions. This enhanced performance in the morning implies that sleep data may be a more potent predictor of morning behaviors than those in the afternoon. Such a pattern might relate to the direct influence of sleep quality on cognitive and emotional states, which likely decreases as the day unfolds. This theory is consistent with the research by Tzischinsky and Shochat [54], which found a strong correlation between sleepiness, sleep-problem behaviors, depressed mood, and the quality of life in adolescents, with a more pronounced effect observed in the morning.

The mentioned factors contributing to the variability in the prediction accuracy of our model can be categorized into two types:

1. The model’s differing performance throughout the day, with higher accuracy observed in the morning compared to the afternoon.
2. The varying predictive success across different residents.

To mitigate the first issue, we propose integrating the sleep monitoring-based model with additional sensing technologies employed in this study through out the daytime, such as Electrodermal Activity (EDA) wristbands and Heart Rate Variability (HRV) monitoring, and vision pipelines. This combined approach aims to develop a more robust and scalable end-to-end predictor for adverse behavior, enhancing its effectiveness beyond morning predictions and updating the likelihood of adverse behavior based on sensory data collected throughout the day.

Addressing the second challenge involves incorporating non-sleep data into our model. This includes analyzing historical patterns of adverse behavior, considering the intensity and duration of such incidents, the history of absenteeism, and medication usage. By integrating these diverse data sources, we aim to refine our model’s predictive capabilities across a broader spectrum of residents.

In future works we also envision focusing on those who did not benefit much from our prediction model. We will explore if their behavior, health conditions, and medications differ from those who responded well. We also plan to improve the accuracy of our labels, which are currently affected by human judgment errors. This will help reduce inconsistencies and noise in our data.

Finally, it is important to highlight the inherent advantages of our proposed end-to-end, privacy-preserving system, which leverages off-body sensing. This methodology offers the potential for minimal intrusiveness when applied, trained, and fine-tuned for the ASD population. It facilitates learning the likelihood of certain behaviors by forming a closed feedback loop, continually updating the system. A key attribute of this system is its adaptability, making it easily integratable into larger healthcare facilities. This integration could significantly enhance monitoring capabilities and resource management in these settings, with additional applications in elopement, falls, and sleep interventions.

## 6 Conclusion

This article presented a novel approach for predicting adverse daytime behaviors in children with ASD by examining the correlation between sleep quality and subsequent daytime behavior. We employed temporally and spatially downsampled footage from low-cost near-infrared cameras recorded during overnight sleep to predict next-day adverse behaviors. An ensemble predictive model was developed to address four scenarios: target-insensitive and target-sensitive predictions for both morning and afternoon behaviors. The research presented in this work provides the first evidence that off-body sensing during sleep can predict daytime adverse behavior in individuals with severe forms of ASD. The described approach is scalable, low-cost, privacy-preserving, and relatively simple to set up and automate. The described system can serve as a novel monitoring tool, allowing caregivers to implement preventative strategies such as reducing levels of environmental stimulation, permitting more frequent breaks, and providing increased access to activities that support emotional regulation. Challenging behaviors can harm self, others, or property and can be socially stigmatizing for the individual; therefore, taking steps to reduce the likelihood of their occurrence is beneficial. This technology could be a valuable resource for caregivers and clinicians who support those with ASD.

## Data Availability

All data produced in the present study are available upon reasonable request to the authors.

## Acknowledgments

This work was supported by the Center for Discovery. YK, PS, AR and GC were partially supported by James M. Cox Foundation and Cox Enterprises, Inc., in support of Emory’s Brain Health Center and Georgia Institute of Technology. GC is partially supported by the National Center for Advancing Translational Sciences of the National Institutes of Health under Award Number UL1TR002378. The content is solely the responsibility of the authors and does not necessarily represent the official views of the National Institutes of Health, the Center for Discovery or the Cox Foundation.

